# Good communication is critical to supporting people living and working with a rare disease: current rare disease support perceived as inadequate

**DOI:** 10.1101/2020.05.22.20110056

**Authors:** Julie McMullan, Ashleen L. Crowe, Caitlin Bailie, Kerry Moore, Lauren S. McMullan, Nahid Shamandi, Helen McAneney, Amy Jayne McKnight

## Abstract

**Background:** Many people living and working with rare diseases describe consistent difficulties accessing appropriate information and support. In this study an evaluation of the awareness of rare diseases, alongside related information and educational resources available for patients, their families and healthcare professionals, was conducted in 2018-2019 using an online survey and semi-structured interviews with rare disease collaborative groups (charities, voluntary and community groups) active across Northern Ireland (NI).

**Results:** Ninety-nine participants engaged with the survey with 31 respondents providing detailed answers. Resources such as information, communication, ‘registries’, online services, training and improvements to support services were queried. Excellent communication is an important factor in delivering good rare disease support. Training for health professionals was also highlighted as an essential element of improving support for those with a rare disease to ensure they approach people with these unique and challenging diseases in an appropriate way. Carers were mentioned several times throughout the study; it is often felt they are overlooked in rare disease research and more support should be in place for them. Current care/support for those with a rare disease was highlighted as inadequate. Nine semi-structured interviews were conducted with rare disease collaborative groups. Reoccurring themes included a need for more effective: information and communication, training for health professionals, online presence, support for carers, and involvement in research.

**Conclusions:** All rare disease collaborative groups agreed that current services for people living and working with a rare disease are not adequate. An important finding to consider in future research within the rare disease field is the inclusion of carers perceptions and experiences in studies. Due to the unique role a carer has in the life of a person with a rare disease it is vital that their voice is heard and their needs are listened to. This research provides insight into the support available for rare diseases across Northern Ireland, highlights unmet needs in service provision, and suggests approaches to improve rare disease support prioritising improved information and communication provision, improved access to services, and tailored support for carers of people with a rare disease.

## Background

In Europe, diseases that affect less than 1 in 2000 individuals are considered ‘rare’ [1]. Collectively rare diseases are a major public health issue affecting approximately 300 million people globally and ~110,000 individuals across Northern Ireland (NI) [2, 3]. The role of healthcare professionals is made more difficult when supporting those with a rare disease due to the limited number of patients with each condition, scarcity of knowledge, uniqueness and complex care of needs [2, 3]. The UK strategy for Rare Diseases emphasised the need for improvement of the coordination of care, diagnosis, treatment and patient empowerment aiming to *‘improve the lives of all those with rare diseases in the UK’* [4, 5].

Many people living and working with rare diseases struggle to access appropriate health and social care, source useful information, or participate in research [2, 6-10]. There is often a lack of appropriate health services, skilled health professionals and effective treatment options [11]. Alongside multiple medical issues that are often associated with a rare disease, it is common for patients, families, and carers to experience psychological stress, loneliness, tiredness, discouragement, unemployment, lack of information and difficulty accessing appropriate health care [11]. This is often made worse by a lack of appropriate peer and community support services [12].

Health and social care professionals frequently experience frustration and distress when caring for individuals with a rare disease as they pose distinct challenges to clinicians working in primary, secondary and tertiary health care settings [13]. Difficulties with diagnosis and management often stem from lack of knowledge, lack of evidence-based information or difficulty accessing information [7, 14, 15]. Because there are approximately 8000 rare diseases, it is unrealistic to provide training for health professionals on every disease [11]. The internet is increasingly used as a source of information and support by families affected by rare disease as a way of gaining information and connecting with others [16].

It has been found that individuals and families affected by a rare disease can benefit greatly from peer support from those facing similar challenges, yet the majority of rare diseases do not have a dedicated support group [17]. Many generic resources exist which can be useful for those living with a rare disease, for example, Global Gene, Rare Disease UK and locally the Northern Ireland Rare Disease Partnership [18-20]. It is interesting to note the difference between the support available for those with a rare disease compared to those with other conditions. For example, those with cancer or heart conditions have access to a wide range of support [21, 22] which receive much greater national funding support than rare disease charities. Gaining an insight into the services rare disease collaborative groups provide and identifying the main services available could be of great benefit to those with a rare disease. Effectively signposting individuals to the most appropriate services would reduce the endless hours of internet searching often undertaken by those with a rare disease and their families. Being aware of the services on offer would also reduce the chance of services being replicated needlessly as well as identifying any gaps in services.

In this study, an evaluation was conducted of the awareness of rare diseases, related information, and educational resources available for patients, their families and healthcare professionals across Northern Ireland. Additionally, there was a focus on how collaborative groups use social media in relation to rare disease(s). An online survey and individual interviews were conducted with representatives from rare disease collaborative support groups as part of the evaluation phase of key priorities for action in the Northern Ireland Rare Diseases Implementation Plan [5].

## Results

### Survey

Ninety-nine respondents participated in the survey with 31 respondents providing detailed answers to questions. The template survey is provided Additional file 1.

#### Collaborative group characteristics

The survey was open to all charities, voluntary and community groups providing services for people living and working with rare diseases across Northern Ireland and anyone from within the group was eligible to answer the survey. Detailed survey responses are provided in Additional file 2. Rare disease collaborative groups were named in the survey to ensure no overlap where multiple representatives may have responded on behalf of a single group; there was no overlap in group names for individual responses. All groups provided services to individuals living and working with rare diseases in Northern Ireland and were established between 1980s to 2017. The majority were registered charities in the UK, 18% were based in Northern Ireland, two were registered in the Republic of Ireland and two were from the United States of America. Groups ranged from providing support in the last two years to more than 50 years experience as a collaborative group. The majority of group representatives completing the survey were White British (60%) or Irish (32%), over the age of 35 (84%), and identified as female (84%). 88% of respondents indicated they were willing to be contacted for follow-up by email, with only two preferring a telephone call.

#### Communication

Ninety-two percent of groups provided or actively supported a facility to put individuals (for example health care professionals, patients, families, carers) in contact with each other on request. While only 14% of groups offer a 24-hour helpline, 41% offer an office-based telephone helpline during the day with 86% of groups offering some form of contact-response option. Ninety-four percent of groups have a dedicated social media representative, while half of groups surveyed had a named information or communication manager, and 68% have a branch or contact person located in Northern Ireland. For many rare disease collaborative groups, improved resources to have ‘better’ helpline services were described as ‘very important’.

As additional figure displays the supportive networking options provide by many groups to help connect people [see Additional file 3]. All groups were interested in collaborating with relevant groups to develop complementary information resources, with a particular focus on opportunities to improve information sharing between community/voluntary and statutory services.

Only one of the groups surveyed does not have a website, the remainder described their website as benefitting anyone affected by a rare disease or is worried about a loved one. Groups differed in terms of when websites were updated, with just four groups providing daily updates and the majority (62%) *ad hoc* as new information is available and personnel resources permit. Two groups have information resources meeting NHS England’s information standard [23], while two have information approved by medical advisors. A variety of social media platforms are used by the groups including Facebook, Twitter, YouTube, LinkedIn, Instagram and Snapchat. Uses of social media within the groups varies but include: sharing information about your specific rare disease (94%), raising public awareness of rare diseases (94%), promoting fundraising campaigns (88%), sharing information about upcoming events (83%), posting information about clinical trials (75%) and sharing recent news (75%). This information is displayed in Additional File 4. Eight groups provide daily updates to social media, with four providing weekly updates, and the remainder updating on an *ad hoc* basis. Three of the groups surveyed do not use social media due to either lack of expertise, privacy concerns and constraints on time.

More detail on social media use by collaborative groups is provided in Additional file 5. 81.3% of groups strongly agree that social media use by rare diseases collaborative groups is beneficial for communication with and support for patients. The majority of those surveyed believed their group should expand its use of social media if time and budget were not issues to be considered. Suggestions made for expansion included: digest news and updates and share across all aspects of the condition, graphic design, educational and awareness raising videos and website development, a daily blog, advice for doctors, nurses, consultants and other health persons to aid understanding of rare conditions and how best to manage them.

#### List of members and registry ideas

95% of groups surveyed hold a list of members with information including: name, address, contact details, type of disease, date of symptoms starting, date of diagnosis, living alone or with carer, family details, mutation types, resource for clinical trials and interest in volunteering. Many groups (36%) struggle with data protection issues in terms of extra work, keeping governance up-to-date, and maintaining contacts, particular those groups that are volunteer led. Although only eight groups hold a disease ‘registry’, with content ranging from simple names and contact details to information on emerging therapies and available clinical trials 77% indicated that they would like one whether held nationally or supported to host themselves. The majority of ‘registries’ were resourced from charitable funds, with one receiving government funding. The distribution of registry data is displayed in Additional file 6.

#### Training

Forty-four percent of groups had developed/co-ordinated training modules for healthcare professionals. Thirty-eight percent of the groups organise training days for healthcare professionals in specific disciplines such as physiotherapists, occupational therapists, hospice staff; some also support members by organising unofficial talks and training for schools, teachers and families. Three groups developed co-ordinated smartphone-based apps including an accident and emergency app for the community, a Patient Buddy app, with another under development.

#### Improving services for those with a rare disease

Various suggestions were made as to how information and communication for rare diseases could be improved inside and outside of the groups and can be viewed in Additional File 7.

When asked how groups would like those living with a rare disease in Northern Ireland to primarily interact with their information resources groups responded with channels such as social media, website, telephone. One group however said they would like live coverage of group meetings to enable them to join remotely. Groups expressed their desire for healthcare professionals to interact with the group’s information resources by joining consortiums, pass on relevant leaflets, as well as the usual channels. Similar methods of communication were suggested for non-clinical professionals and for other groups.

### Interviews

Nine semi-structured 1-1 interviews were conducted with rare disease collaborative groups based in Northern Ireland lasting 20-80 minutes. The interview participants were either in paid employment or a held a voluntary role within a rare disease support group. The duties of these individuals varied from supporting patients, holding meetings, organising conferences, advisory roles, signposting, advocacy and raising awareness. Half of the collaborative groups interviewed has a focus on a rare disease that is genetic although most participants acknowledged having a limited knowledge of genetics and relied on specialists to provide them with this information when required.

The enthusiasm the participants had for the collaborative group they volunteer/work for and the individuals they support was evident from the interviews. They were passionate when they spoke and often failed to recognise the amazing work they do and the difference they are making. Each person felt they would like to do more to help and put forward several suggestions for how care for rare disease patients might be improved.

The participants’ identities have been protected by using the codes P1-P9, three of these participants also completed the survey. Thematic analysis [24] was used to analyse the data; six themes emerged. Findings from the interviews are outlined thematically below with direct quotes used for representative samples.

#### Theme 1: Excellent communication is key

This theme emerged from participants’ views that effective communication around rare disease creates an understanding and improves care.

> *…” communication, I always define it as creating understanding, that’s what it is, communication is the key to everything.” (P7)*

Throughout the interviews it became apparent that communication around rare disease involves several groups: those with a rare disease, carers, family members, health care professionals and the voluntary sector. With several collaborative groups working in Northern Ireland to support those with a rare disease, communication is vital to ensure that the work being carried out is complimentary and is an effective use of resources.

> *…” you don’t want to duplicate the work either that somebody else has, so it’s that thing about networking and communication” (P3)*

The majority of the groups interviewed work with other rare disease groups and feel they have much to learn from one another. The groups provide links to other rare disease organisations via their website and find networking opportunities to be extremely useful, at times even raising their group’s profile.

> *…”networking… to try and map what research is going on and they all communicate with each other”. (P5)*

Most groups have a helpline that ranges from an answering service on a mobile phone to a 9 am – 5 pm manned telephone. The helplines are mainly used for supporting those who are newly diagnosed when they feel they have nowhere to turn or to signpost people to other means of help. Most groups reported that initial contact from rare disease patients was normally *via* social media or email rather than the phone line.

Difficulties were highlighted by many groups regarding meeting to communicate in person with someone living with a rare disease, for example at a conference or support group. This is thought to be due to the nature of the disease and geographical boundaries that can inhibit an individual’s chance to attend.

> *“Attendance at these meetings is very poor but that’s the nature of the condition, you’re either very ill and you can’t get to the meeting or maybe you physically can’t get to the meeting or you’re well and you don’t want to be reminded that you have this illness”. (P2)*.
>
> *…” social media…. primary means of communication to the outside world.” (P1)*

#### Theme 2: Online presence

> *…” there’s quite a lot of interaction between patients…” (P5)*

Several uses of social media were highlighted by the groups interviewed including: advertising surveys, links with other charities, research updates, highlighting events/fundraising and posting photos. The instantaneous benefits of social media were acknowledged by the groups. Managing the platforms and the possibility of missing individuals who do not use the internet were raised as concerns of using social media. Invaluable support and mentoring from those experiencing something similar was also referred to as a positive outcome of using social media in relation to rare disease. One group explained they sometimes have a professional on the Facebook page for an hour to take queries and then an administrator will upload the answers.

> *.. ” when we have the questions and answers with the professionals very well, you know people have put aside that hour in their evening and they’re willing to sit in and listen and put their questions…” (P3)*

A number of uses of the group’s websites were highlighted including: making initial contact with the group, guidance and advance regarding clinical management, displaying factsheets of key symptoms; practical management; red flags; lists of the conditions which are considered rare so it is clear who the group can help, information about research; information for health professionals; information about the support groups; link to other forums; hub – local resources, generic information for example blue badge guidance, links to specific support within hospitals. For most groups the general aim to of the websites is to include content that will be relevant for anyone who has an interest in rare diseases whether newly diagnosed or wanting to know what is currently happening.

> *…” Predominantly the reason is awareness raising but also to provide a point of contact so they can actually make contact…” (P7)*
>
> *..”anybody who has any interest in rare disease issues will find something useful there whether it’s someone who’s recently been diagnosed and has no idea what’s going on where to go or who to talk to or whether it’s somebody who has a bit of an interest and wants to know what’s happening with other people, what level of engagement there is”. (P6)*

#### Theme 3: Carers

Participants were asked directly during interview if they provide support for carers. Most groups said that although carers are welcome to attend events they organise, they do not host anything specifically for carers. The quote below illustrates this:

> *“We’re very, very conscious about carers or family members because they go through the same kind of disease or that the patients experience so we do, whenever we plan anything we always take into account family members and carers. I mean there’s not a dedicated network for family members but it’s something that we are looking into”. (P8)*

The participant below explained that carers are involved with such a highly intensive care routine that they struggle to find time to attend support groups.

> …. *“carers are really stretched, they don’t have a lot of free time, even if they wanted to come to support groups still there isn’t the respite care that will cover them. ...” (P5)*

Other participants were in agreement that the needs of the carers are as important as the person with the rare disease and they should not be overlooked. One participant said that their group previously tried to organise a carers only workshop at a conference but it was unsuccessful. They stressed how they feel it is important for carers to get time away to allow them to off load and discuss any issues they are facing.

> *…” I’d really like at times the carers just to talk without the patients there you know no harm to them or anything like that but we need to vent our issues.” (P2)*

In most cases the groups stated that the initial contact with the group comes from a family member or a carer who are reaching out on behalf of the person they are caring for. This indicates the complex role and unique relationship that the person with the rare disease has with their carer.

#### Theme 4: Training health professionals

> *“We do lots of training for health professionals. I would say probably at least every month we have a training session somewhere for health care professionals.” (P5)*

More than half of the groups interviewed have involvement / training experiences with health professionals, those that do not said this is due to struggling to get professionals on board. The training discussed during interviews included teaching at Queen’s University, Belfast (QUB) presentations to health professionals, e-learning modules, questions and answers with health professionals on social media and encouraging health professionals to attend the conferences. One group explained that they usually have someone attending their clinic once a month and then they encourage them or even the genetic trainees to do the BMJ module relating to their specific disease.

> *…” we contribute to medical education…” (P6)*
>
> *…” we have e-learning modules, the first 2 that were launched were for GPs and physiotherapists and they were really successful”… (P9)*

Another group said they provide a variety of training for health professionals, from large regional or national events with lots of multidisciplinary teams, to events on a much smaller scale with local teams. They also hold a health care professionals day each year to raise their awareness and help them to realise that although a rare disease is often considered ‘specialised’ the ongoing processes and problems it causes are transferable across lots of conditions and so the issue is about how you support the person as the emotional impact is the same. Another group felt that patients have the *‘lived experience’*, therefore we can all learn from each other. They said that patients can lose confidence in the health service when they repeatedly hear, *‘I’ve never heard of this’*.

#### Theme 5: Involvement with research

Three of the groups interviewed have had involvement with research. One group recognised the fact that not everyone wants to know everything about their rare condition but that others are extremely interested in the latest research in their area and aim to know as much as they can about it.

> *…’’Twitter feed and I post any relevant research I’d post on that cause I don’t want to drown the Facebook group with research because not every parent is on that level”… (P1)*

Several groups use Twitter to promote research articles relating to their specific area. Groups acknowledged the importance of research into rare disease, highlighting the need for engagement with specialists to address the patient’s needs, and the need for awareness raising of research to show the importance of getting patients involved.

> *“I started to realise that if we get involved at the very beginning, if the idea comes from us about what we need then there’s no this waste of, like so money has been wasted, so much money has been put in to getting 1 drug on the market you know but you really want that to be something that people need that’s going to be better than what already exists”. (P2)*

Networking and collaborating with other countries was also referred to as an important aspect of research into rare diseases. Groups acknowledged the difficulties that can be encountered when trying to get the manpower to look for grants or funding and the money that can be wasted by not involving patients from the beginning.

> *…“got that networking going em, to try and map what research is going on and they all communicate with each other so if you get a new young clever person who’s interested in doing a bit of research, there’s a bit of guidance as to what needs to be done next, where we’re at and what needs to be done next”. (P5)*

#### Theme 6: ‘Not adequate’

All of the groups interviewed were in agreement that the services for rare disease patients in Northern Ireland (and further afield) are not adequate. Figure 5 outlines some of the improvements that were suggested by survey respondents.

> *…’’definitely there could be more done”. (P9)*

Suggested improvements included: funding organisations such as NIRDP; partnership between statutory and voluntary services, referrals to groups from health service; share information, network of people to learn from each other. Some groups highlighted the difficulties newly diagnosed patients face and how work is needed to make people aware of the kind of help that is available.

> *“…just with a new diagnosis it’s just that initial feeling of where to go but I think once somebody gets into and starts networking, I think the realisation is that there are things out there”. (P3)*

Groups explained that having health professionals on board with the work the groups are doing helps to promote their work, raise awareness and achieve more.

> *“You need a champion and that came across very, very strongly at the last rare disease [meeting] in March, you know the all Ireland conference was the groups that had their champions, they were the ones who actually achieved more and you need somebody behind who is totally invested who wants to push things forward”. (P4)*

Other suggestions included: connecting with centres of expertise; educating people; better communication; not duplicating work; involving more experienced people with the social media; better co-ordination of care; developing an information hub; making people aware that their condition is ‘rare’; discovering what rare disease patients have in common; listen to patients; smoother access to health care; more support needed for carers (perhaps in a way that can be accessed remotely). Often respite is not provided and therefore they cannot attend organised events; awareness raising is needed.

> *“A big gap continues to be the coordination of care. It continues to be people know knowing where to go, people, healthcare professionals not having confidence in their own abilities.” (P6)*
>
> *“…there just isn’t enough resource out there for the sorts of needs that people have…” (P5)*

Several ‘novel’ ideas were suggested by the groups as ways to improve or enhance the services already available for those living with a rare disease. These included an app to enable patients to better manage their condition, an online map which links to health professionals/support in a specific geographical area, training on practical issues that are relevant across many rare diseases, the opportunity to join conferences/events remotely and a ‘rare disease community’. An interesting point was made by one group that individuals may not know they have a ‘rare disease’ as this terminology may not have been used to describe their condition making it more difficult for them to find appropriate support.

## Discussion

This study aimed to evaluate rare disease resources in Northern Ireland and to identify important issues for local development. Little research has been conducted to date reviewing dedicated support available for people with a rare disease and to date no one has reviewed group resources for those with a rare disease. This is particularly challenging where no disease-specific support group exists for individuals with a named rare disease, or those who are waiting on a diagnosis

### Communication is an important factor to consider when developing rare disease support

In agreement with previous research, communication was found to be an important factor when considering support for those with a rare disease [7]. Pelentsov *et al*. (2016) acknowledged how challenging communication can be for those with a rare disease, particularly those who are newly diagnosed [25]. Improved communication between General Practitioners (GP) and other healthcare professionals would help better manage care for patients with rare disease(s). Rare disease registries are an essential tool to measure the number of people affected by a rare disease(s) and thus contribute to local evidence-based service planning. Such registries also improve knowledge and monitor interventions for rare diseases and provide longer-term understanding for a specific disease [26, 27]. The majority of groups in the current study expressed their desire for a rare disease registry to be established in NI. The disconnect between the health service and groups was mentioned and it was suggested that a registry may help the health service to be more responsive to rare disease patient’s needs. The development of rare disease registries is growing which is remarkable given the scarcity of patients [28]. A rare disease registry could be a powerful tool for rare disease patients, enabling long-term outcomes to be measured and ultimately improving care [29].

Communication methods need to be considered carefully with particular attention given to equity of access to good communication tools. All of the groups surveyed or interviewed offer those with a rare disease various means of communicating from helplines, to social media and websites. Collaborative groups acknowledged the difficulties that are often experienced when trying to organise face to face meetings. This is largely due to the nature of rare diseases and the unpredictable symptoms experienced on a planned date. It seems there is a move away from this type of support towards online platforms and remote access to in-person / virtual meetings.

Social isolation is a consistently recurring theme for those with a rare disease, and the internet offers a practical solution particularly in rural areas [7, 30]. Many of those living with a rare disease find belonging by being part of an online support group or blog [31]. Although the merits of social media and online support are many, it is important to consider the reality that many people do not regularly access online services and so there is a chance that people could be missed, as raised by participant 7. The most common social media platform used by groups was Facebook, followed by Twitter. As in agreement with Lasker *et al*. (2005) social media was found to be used for ‘sharing information about the specific rare disease’ and ‘communication with other organisations’ [17].

100% of groups said they would be interested in collaborating with relevant groups to develop complementary information resources. This is an idea which should be developed to bring services together and provide a clear pathway for rare disease patients. Listening to patients was also raised as an issue in this study by collaborative groups as many patients do not feel the communication between themselves and health professionals is good enough. Jeppeson *et al*. (2014) also found this, stating that gaining an insight into how someone copes with the day to day life of living with a rare disease would enable a deeper understanding and encourage more sympathetic communication [12]. Rare disease patients often experience a shift in their relationship with health professionals as they and their carers take on the role as the ‘experts’ [32]. Budych *et al*. (2012) have shown improved outcomes will result from care providers recognising and acknowledging patient’s expertise and being willing to partner with them in the decision making around treatment [33]. Many collaborative groups discussed how having health professionals on board with their work would help to champion and promote what they do.

The NIRDP set out four priorities within their vision as a charity. The first is ‘Connecting’: Stronger Together; Outreach to the rare disease community in Northern Ireland; Tackling disadvantage and inequality; Reaching out to those living and working with rare disease elsewhere in the UK and Ireland, and across the world [20]. From this element of the charities vision it is apparent that communication is an important aspect of the work they do within Northern Ireland which supports the need identified within this project.

### Training

Effectively supporting those with a rare disease requires training. The majority of groups who were part of this study are involved with training health care professionals with several having developed/co-ordinated training modules. Although the Northern Ireland Rare Disease Implementation Plan (2015) attempts to support both GPs and rare disease patients there remains a lack of information among the GP populations [5, 6]. Greulich *et al*. (2013) among others highlighted the lack of knowledge of general practitioners on the topic of rare diseases and suggested a web-based module might increase their knowledge of the specific disease, generic complications (e.g. providing respiratory support is necessary for multiple rare diseases) and improve patient care [34]. Multiple groups explained that they organise training days for healthcare professionals and specific disciplines such as physiotherapists, occupational therapists, hospice staff and unofficial talks and training for schools, teachers and families. A central online resource listing such events would be beneficial to many people. The UK strategy for Rare Diseases emphasised the need for improvement of the coordination of care, diagnosis, treatment and patient empowerment [4].

Although plans and initiatives are in place, the GP community still do not feel adequately equipped to deal with rare disease with many frequently consulting online databases such as Google, Orphanet and Online Mendelian Inheritance in Man (OMIM) [35]. Fernández Avellaneda *et al*. (2006), investigated the need for rare disease training in primary care [36]. Difficulties arose in understanding the importance and magnitude of rare diseases, supporting the findings of the current study that further training is required. Ramalle-Gomara *et al*. (2014) conducted a survey with 234 students and found that only around 25% of them knew the definition of rare disease and an orphan drug, again highlighting the need for training and awareness raising [37, 38]. Further training is needed to appropriately equip people to support those living with a rare disease [5].

### Carers

Carers for individuals with a rare disease were frequently discussed - the main issue being that carers are regularly overlooked and yet they often need support just as much as the person with the rare disease that they are caring for. Living with a rare disease often involves many appointments and can include travelling long distances to see specialists or receive treatments and so daily routine can be disrupted for families [39]. This is particularly challenging in the era of COVID-19 as all hotels and accommodation options within the UK are closed to everyone except NHS staff; families who normally stay overnight for complex and / or tiring treatments are unable to do so, thus compromising care and raising inequalities for those living geographically further from treatment centres [40]. For parents of children with a rare disease, balancing daily life with caring is a frequent problem and often carers neglect their own needs [41]. It is therefore important as this study suggests that strategies and support are put in place for carers. Little support is currently provided specifically for carers, it is important that these individuals are not overlooked.

Those living with a rare disease face considerable barriers when accessing appropriate care including delayed diagnosis and limited or non-existing treatment options [42]. The rare disease patient community has played a critical role, in response to these challenges, elevating patient voice and mobilising legislation to support the development of programs that address the needs of patients with rare diseases [42]. For many of the parents of a child with a rare disease, the burden of care spans many years and involves a lifetime commitment. Parents will require specialist health literacy, care giving skills and resources beyond those normally required by parents in general [25].

Rare diseases result in a wide variety of healthcare needs [43]. Rare diseases significantly impact the economic, psychosocial, and physical well-being of individuals and their family members [39]. Individuals with rare diseases and their family members often have limited evidence-based information to guide their decisions about disease management and symptom relief [44, 45]. Further, the inherent uncertainty that comes with having a rare disease, including delays in diagnosis and a lack of knowledge about current and future care needs, impact access to services and management of the rare disease [46-48]. Research on the experience of having a rare disease indicates that care and services needs are often not determined simply by the severity of the health condition, but rather are a combination of poor quality of care and barriers to access [47, 49]. Baumbusch *et al*. (2019) concluded that informal peer support from other parents of children with rare diseases were a key resource for parents when navigating the health care system. Social media was seen as the best way for these people to connect, giving the best availability [43]. A research study focusing on carers alone would be beneficial for this cohort to identify their needs and how they can be best supported in this challenging role.

### Current care/support for those with a rare disease is not adequate

All collaborative groups interviewed were in agreement that current services for rare disease patients in Northern Ireland and further afield are not adequate. Ensuring funding is made available to rare disease collaborative groups was an important factor to consider when developing future services. Many charities and support services are available to help people with various diseases however ‘rare disease’ support is lacking in comparison to cancer and heart patients for example. Pelentsov *et al*. (2015) outlined the benefits of support groups for those with similar diseases [25]. In agreement with this study, Taruscio *et al*. (2014) suggested that an accessible one-stop-shop source of information and support should be an important goal for patients with a rare disease [50]. A reference network ‘hub’ in Northern Ireland would bring many benefits.

There is a requirement for further research and development in rare diseases in order to meet patients’ medical needs [51]. Within Northern Ireland, the NIRDP has made a commitment to the rare disease community stating that no one should be disadvantaged because of the rarity of their condition. NIRDP aim to innovate by developing and implementing improved methods of managing and treating rare diseases; improving the quality of life for those affected by rare diseases; and increasing the efficiency and effectiveness of care and support [20]. Some of the ideas for developing and improving rare disease support suggested by the collaborative support groups included new and innovative ideas as well as some things that are already being done by other groups: apps for patients to manage their conditions; online map to link to health professionals/support by geographical area; a hub to co-ordinate services. It is interesting that access to information for therapies, aids, grants and clinical trials for example seems to differ a lot from the help and support available to non-disease-specific RD charities compared to non-disease specific cancer charities. Despite this, many countries have made great progress in improving processes in the rare disease landscape with patient advocacy playing an increasing role in shaping discussions and driving the agenda [42]

#### Strengths and limitations

This study explored an area of very limited published research. The use of an online survey made it extremely accessible for a wide audience to access remotely. The data collection procedures included surveys and semi-structured interviews which were promoted online via the NIRDP and QUB; other methods of dissemination may have helped to generate more respondents/participants. In future research print media and local radio should be considered as options to enhance recruitment. To be eligible for interview participants had to be English speakers, it is possible that this may have introduced bias to the sample in that potential participants may have been excluded, but due to the language skills of the research team this was an essential criterion.

## Conclusion

This study has provided the voluntary sector with an opportunity to share their views and experiences of the current support available for rare diseases. New insights are added to the existing body of knowledge on rare disease, highlighting the need for future research including carers and more training for health professionals regarding communication for rare disease. The surveys and interviews conducted with rare disease collaborative groups have provided insight into the support available for rare disease patients in Northern Ireland and have identified the gaps in services. From this information it has become obvious that many valuable services exist for rare disease patients and that those providing these are extremely passionate and dedicated to the work that they do. A clear area to target would be communication, there appears to be a disconnect between statutory and voluntary services and a better link would improve the care provided for rare disease patients. Access to services for those who are geographically more distant or are not well enough to attend events is also an area worth targeting. Perhaps with the increasing use of social media there would be a way to enable this group to attend and interact with events in a remote way. The current challenges associated with social distancing to protect individuals who are most clinically vulnerable from COVID-19 provides a valuable opportunity to review the benefits and limitations of more remote activities in the coming year.

Communication is key for rare disease support groups, it is important that communication is both accessible to all and is reciprocated from health professionals. All groups were interested in collaborating and therefore improving lines of communication and enabling opportunities for collaboration could help greatly in developing and improving services. There were also pleas for a local rare disease ‘registry’. Social media was used widely within the groups that were part of this study although it was noted that various difficulties can arise when trying to maintain these accounts. Every group made suggestions as to how they feel services and support could be improved for those with a rare disease, many of these ideas centred on communication as well as increasing resources. The fact that carers are not often catered for specifically by these groups is something needs to be addressed as it was highlighted that in most cases it is the family member or carer who makes the initial contact with the group. The struggle to get health professionals on board was acknowledged. There was an agreed consensus that the services for rare disease patients in Northern Ireland are not adequate.

Further research should be conducted with rare disease collaborative groups, specifically it would interesting to get the views of those accessing these services. Research focusing on carers would also be useful as to date no research studies focus solely on them. Interestingly, many studies have been conducted on carers for cancer [52, 53, 54]. The lack of research suggests that a difference exists between cancer caring support options and those for rare disease patients. Families living with rare diseases have traditionally received little attention from health authorities, clinicians and researchers [44]. Given the unique and close relationship carers often have with rare disease patient(s) it is crucial that they are included in future research.

Locally and internationally it is important to build upon existing support and move towards a brighter future for those with a rare disease. Additional file 8 shows the recommendations to inform health care policy and practice

## Methods

In this study, an evaluation was conducted of the awareness of rare diseases, related information, and educational resources available for patients, their families and healthcare professionals across Northern Ireland. A primarily qualitative approach was chosen for this study as the aim was to explore the current services provided by rare disease collaborative groups across Northern Ireland by gaining the views and perceptions of individuals who work/volunteer for them. Qualitative research aims to understand behaviour and beliefs, identify processes and to understand the context of people’s experiences [55]. In this study data were collected from a relatively small population group but findings may be applicable for a wider population due to the common challenges and specific needs of those with a rare disease.

The study had two stages:

- *Stage 1:* Online survey with rare disease collaborative groups in Northern Ireland.
- *Stage 2:* Semi-structured interviews with rare disease collaborative groups in Northern Ireland.

The survey and interviews were used to help evaluate awareness of rare diseases, alongside related information and educational resources available for patients, their families and healthcare professionals in Northern Ireland. They were conducted as part of the evaluation phase of key priorities for action in the *Northern Ireland Rare Disease Implementation Plan* [5]. Results were collated anonymously and used to help improve the accessibility of resources and address the needs of individuals affected by rare disease(s) in Northern Ireland. Through the surveys and interviews we wanted to locate existing resources as well as identify gaps where the development of further resources would be appropriate.

### Stage 1: Online surveys

A survey was constructed using an iterative approach with input from patient and healthcare professional representatives from the Northern Ireland Rare Disease Partnership (NIRDP, the national charity for rare diseases in Northern Ireland) [20] and was uploaded onto SmartSurvey [56]. The survey was also promoted on by the NIRDP (Twitter, @NI_RDP, 1,167 followers; Facebook, @NIRDPNews, 952 members) and QUB through their websites and associated social media platforms beginning early 2018. Further dissemination occurred by individuals, member charities, and voluntary groups; there was not a mechanism available to track the reach of this extended dissemination. Collaborative rare disease groups that were member charities of NIRDP or had engaged with previous research by our team [57] with consent for recontact were contacted directly to alert them to the survey and interview invitations. Potential participants who expressed an interest were then sent an email with a cover letter (including a link to the survey) and a participant information sheet, which informed them of the purpose of the study and guidance on how to complete the survey.

The survey was opened early 2018 for a period of 3 months. Each section in the survey contained closed and open-ended questions and were displayed with the option to save and return at a later date. Question four of the survey asked participants for permission to be contacted for follow-up questions *via* an interview which aided recruitment for the next stage of the study. SmartSurvey was used to generate tables and charts to add demographic and statistical information to this study [56].

### Interviews

Semi-structured interviews were chosen for this study as they allow the interviewer to conduct the conversation using their own words and are most commonly associated with the collection of qualitative social data when the researcher is interested in people’s experience [58, 59]. An interview schedule was used which helps the researcher to conduct the interview by outlining what will be explored while allowing flexibility for details that are unique to each participant [60]. As there have been few previous qualitative studies undertaken in this area, the interview schedule was designed specifically for this study with the aim and objectives guiding the questions with input from the NIRDP.

A total of 36 emails were sent to rare disease collaborative groups across Northern Ireland with a covering letter and a participant information sheet which provided details of the purpose of the study and explained what they would have to do should they agree to take part. Reminder emails were sent out one week after the initial email had been sent.

A total of nine collaborative groups agreed to be interviewed (25% of those contacted) within the timeframe of this study. Data collection for interviews took place between July 2018 and May 2019 in a neutral location. Informed consent was obtained prior to each interview taking place, which included permission to record the interview on a digital recorder. The interview schedule provided a loose framework to encourage ideas from the participants. Probes were used in response to the information provided by participants.

Each interview was transcribed verbatim by the interviewing researchers (JM & AC) as soon as possible to maximise the accuracy of the transcription process; interview transcripts were uploaded to NVivo12 software for analysis [61] to describe, discuss, evaluate and explain the content and characteristics of the data that has been collected [62]. Working with qualitative data involves gaining an understanding of the words, stories, accounts and explanations of the participants [63]. Comparing the data from each transcript allowed for meanings to be explored within the data and for the identification of relationships between different parts of the data and for the provision of explanations for the similarities or differences found [64]. The data was analysed using Thematic Analysis [24].

## Data Availability

The datasets generated and/or analysed during the current study are not publicly available due as they contain potentially identifiable information but are available from the corresponding author on reasonable request

List of abbreviations
ECHO Extenasion for Community Healthcare Outcomes
GP General Practitioner
HCP Health care personnel
NI Northern Ireland
NIRDP Northern Ireland Rare Disease Partnership
OMIM Online Mendelian Inheritance in Man
QUB Queen’s University, Belfast
RD Rare disease
UK United Kingdom

## Declarations

- **Ethics approval and consent to participate:** This study has ethical approval from QUB’s School of Medicine, Dentistry and Biomedical Sciences research ethics committee, Northern Ireland. Consent to take part in the survey was explicitly given by all participants by actively ticking a ‘consent box’ before they could proceed with the online survey. Consent for the interviews was collected *via* a hard copy form which was signed by the participant.
- **Consent for publication:** Not applicable.
- **Availability of data and material:** The datasets generated and/or analysed during the current study are not publicly available due as they contain potentially identifiable information but are available from the corresponding author on reasonable request.
- **Competing interests:** The authors have no competing interests to declare. AJM is a former board member and HM an existing board member of the NIRDP.
- **Funding:** JM was supported by an award from the NI Public Health Agency and the Medical Research Council – Northern Ireland Executive support of the Northern Ireland Genomic Medicine Centre though Belfast Health and Social Care Trust. AC and CB are recipients of PhD studentships from the Department for the Economy NI. KM is supported by an award from the Rank Foundation.
- **Authors’ contributions:** AJM conceived of the project, designed the survey with input from patients, healthcare professionals and voluntary groups, and drafted the manuscript. JM collected the survey and interview data, analysed the data, and drafted the manuscript. AC, CB, LM and NS helped with data collection. All authors contributed to data interpretation, manuscript revision and agreed the final version for submission.
- **Acknowledgements:** We thank all individuals and groups who participated in this study. We also thank NIRDP (www.nirdp.org.uk) and QUB (https://www.qub.ac.uk/research-centres/CentreforPublicHealth/) for helping promote the study.

## Additional files

*Additional file 1*

- Survey template
- PDF

*Additional file 2*

- Detailed survey responses
- MS Word

*Additional file 3*

- Supportive networking options provided by rare disease collaborative groups to help connect people
- MS Word

*Additional file 4*

- Top purposes of social media platforms and most popular information offered on websites.
- MS Word

*Additional file 5*

- Social media use by rare disease collaborative groups
- MS Word

*Additional file 6*

- Distribution of ‘registry’ data
- MS Word

*Additional file 7*

- How information and communication could be improved within and beyond rare disease collaborative groups.
- MS Word

*Additional file 8*

- Recommendations to inform health care policy and practice
- MS Word

## References

1. European Union. Regulation (EC) N°141/2000 of the European Parliament and of the Council of 16 December 1999 on orphan medicinal products. http://eurlex.europa.eu/LexUriServ/LexUriServ.do?uri=OJ:L:2000:018:0001:0005:EN:PDF. Accessed 24 April 2020.

2. McKnight A.J., McMullan J, Walker R, & Collins C., Communications and Information Review: NI Report for Rare Diseases, (2020, Jan 31).

3. McMullan J, Crowe A, Kerr K, Bailie C, McAneney H, McLaughlin F, Collins C, McKnight A.J. Perspectives on a NI Rare Disease Registry, (2020, Jan 31).

4. Department of Health. The UK Strategy for Rare Diseases. (2013). Available online at https://assets.publishing.service.gov.uk/government/uploads/system/uploads/attachment dat a/file/260562/UKStrategyforRareDiseases.pdf. Accessed May 1, 2020.

5. Providing High Quality Care for people affected by Rare Diseases-The Northern Ireland Implementation Plan for Rare Diseases. 2015. https://www.health-ni.gov.uk/sites/default/files/publications/dhssps/ni-rare-diseases-implementation-plan-oct-2015.pdf. Accessed 3 Feb 2020.

6. Crowe A. McAneney H., Morrison P., Cupples M. & McKnight, A, 2020., A quick reference guide for rare disease: supporting rare disease management in general practice, British Journal of General Practice. 70, 694: 260–261.

7. Crowe A., McKnight A. & McAneney, H.m Communication needs for individuals with rare diseases within and around the healthcare system of Northern Ireland, In: Frontiers in public health. 21 Aug 2019.

8. McAneney H., Crowe A., Millar J., Kerr K., McMullan J., & McKnight A. J., Improving rare disease identification and coordinating health and social care priorities. European Journal of Human Genetics, 27, 2019 [P19.39C].

9. Crowe A., McAneney H., McMullan J., & McKnight A. Improving communication for individuals with a rare condition. European Journal of Human Genetics, 2019; 27, 696–697.

10. McKnight A. J. Recommendation for a Collaborative Centre of Expertise for Rare Diseases in Northern Ireland (CERDNI). https://pure.qub.ac.uk/en/publications/recommendation-for-a-collaborative-centre-of-expertise-for-rare-d (2013, Sep 30).

11. Elliott EJ., Zuryski YA., Rare diseases are a ‘common’ problem for clinicians, Australian Family Physician; 2015; 44(9):630–633.

12. Jeppesen J., Rahbek J., Gredal O., and Hansen, H.P. How narrative journalistic stories can communicate the individual’s challenges of daily living with Amyotrophic Lateral Sclerosis, Patient. 2014; 8: 41–9.

13. Lopes M.T., Koch V.H., Sarrubbi-Junior V, Gallo P.R., Carneiro-Sampaio M., Difficulties in the diagnosis and treatment of rare diseases according to the perceptions of patients, relatives and health care professionals, Clinics. 2018; 73: e68

14. Roo M., López Martin E., Wilkinson M.D. (2017) Preparing Data at the Source to Foster Interoperability across Rare Disease Resources. In: Posada de la Paz M., Taruscio D., Groft S. (eds) Rare Diseases Epidemiology: Update and Overview. Advances in Experimental Medicine and Biology, vol 1031. Springer, Cham18 Nat Rev Genet. 2018 May;19(5):253–268. doi: 10.1038/nrg.2017.116. Epub 2018 Feb 5 Paediatric genomics: diagnosing rare disease in children. Wright CF^1^, FitzPatrick DR^2^, Firth HV^3^4.

15. Eurodis, Rare diseases Europe. https://www.eurordis.org/ Accessed 30 April 2020.

16. Beall J, Internet Resources in Rare Diseases, Health Care on the Internet, 2001 5: 4.

17. Lasker JN, Sogolow ED, Sharim RR The Role of an Online Community for People With a Rare Disease: Content Analysis of Messages Posted on a Primary Biliary Cirrhosis Mailinglist J Med Internet Res 2005;7(1):e10

18. Global Genes, Allies in Rare Disease. www.globalgenes.org Accessed 1 May 2020.

19. Rare Disease UK. https://www.raredisease.org.uk/ Accessed 1 May 2020.

20. Northern Ireland Rare Disease Partnership. https://www.nirdp.org.uk Accessed 24 April 2020.

21. Cancer Research UK. www.cancerresearchuk.org. Accessed 25 April 2020.

22. British Heart Foundation. https://www.bhf.org.uk/ Accessed 25 April 2020.

23. NHS England https://www.england.nhs.uk/tis/) Accessed 23 April 2020

24. Braun V. and Clarke V. Using thematic analysis in psychology. Qualitative Research in Psychology, 2006. 3 (2), 77.

25. Pelentsov LJ., Laws TA., Esterman AJ., The supportive care needs of parents caring for a child with a rare disease: A scoping review. Disabil Health J. 2015; 8(4):475–91.

26. Bellgard M., Beroud C., Parkinson K., Harris T., Ayme S., Baynam G., Weeramanthri T., Dawkins H. and Hunter A., Dispelling myths about rare disease registry system development, Source Code for Biology and Medicine, 2013, 8: 21.

27. Bilton D., Caine N., Cunningham S., Simmonds N.J., Costgriff R. and Carr S.B., Use of a rare disease patient registry in long-term post-authorisation drug studies: a model for collaboration with industry, The Lancet Respiratory Medicin, 2018 6: (7); 495–496.

28. Kodra Y., Weinbach J., Posada-de-la-Paz M., Coi A., Lydie Lemonnier S., van Enckevort D., Roos M., Jacobsen A., Cornet R., Faisal Ahmed S., Bros-Facer V., Popa V., Van Meel M., Renault D., von Gizycki R., Santoro M., Landais P., Torrer P, Carta C., Mascalzoni D., Gainotti S., Lopez. E., Ambrosini. A, Müller H., Reis R., Bianchi F., Rubinstein Y.R., Lochmüller H., Taruscio, D., Recommendations for Improving the Quality of Rare Disease Registries, International Journal of Environmental Research and Public Health, 2018;15, 1644.

29. Lacasse Y, Krishnan JA, Maltais F, Ekström M. Patient registries for home oxygen research and evaluation. Int J Chron Obstruct Pulmon Dis. 2019;14:1299–1304 https://doi-org.queens.ezp1.qub.ac.uk/10.2147/COPD.S204391).

30. McKnight, A.J. (2015); Living every day with a rare disease.

31. Glenn AD., Using Online Health Communication to Manage Chronic Sorrow: Mothers of Children with Rare Diseases Speak Journal of Pediatric Nursing, 2015; 30, 17–24.

32. Badiu C., Bonomi M., Borshchevsky I., Cools M., Craen M., Ghervan C., Hauschild M., Hershkovitz E., Hrabovszky E., Juul A., Kim S-H., Kumanov P., Lecumberri B., Lemos M.C., Neocleous V., Niedziela M., Djurdjevic SP., Persani L., Phan-Hug F., Pignatell D., Pitteloud N., Popovic V., Quinton R., Skordis N., Smith N., Stefanija MA., Xu C., Young J., Dwyer AA., Developing and evaluating rare disease educational materials co-created by expert clinicians and patients: the paradigm of congenital hypogonadotropic hypogonadism, Orphanet Journal of Rare Diseases. 2017; 12: 57.

33. Budych K, Helms TM, Schultz C, How do patients with rare diseases experience the medical encounter? Exploring role behavior and its impact on patient-physician interaction, Health Policy. 2012; 105(2–3):154-164.

34. Greulich, T., Ottaviani, S., Bals, R., Lepper, P.M., Vogelmeier, C., Luisetti, M., Alpha1-antitrypsin deficiency - Diagnostic testing and disease awareness in Germany and Italy. Respir Med. 2013; 107: 1400–8.

35. Stoller JK., The Challenge of Rare Diseases, Chest. 2018; 153(6):1309–14.

36. Avellaneda Fernández A., Izquierdo Martínez M., Luengo Gómez S., Arenas Martín J., Ramón JR., Need for primary care training in rare diseases. Aten primaria. 2006; 38(6):345–8.

37. Ramalle-Gómara E., Ruiz E., Quiñones C., Andrés S., Iruzubieta J., Gil-de-Gómez J., General knowledge and opinion of future health care and non-health care professionals on rare diseases., J Eval Clin Pract. 2015; 21: 2–198.

38. Ramalle-Gómara, E., Domínguez-Garrido, E., Gómez-Eguílaz, M. et al. Education and information needs for physicians about rare diseases in Spain. Orphanet J Rare Dis 15, 18 (2020). https://doi.org/10.1186/s13023-019-1285-0

39. Zurynski Y., Frith K., Leonard H., Elliott E., Rare childhood diseases: how should we respond?, Arch Dis Child. 2008;1;93(12):1071.

40. Covid19 Advice for accommodation providers, Available online at: https://www.gov.uk/guidance/covid-19-advice-for-accommodation-providers. Accessed 14 May 2020.

41. Etchegary H., Healthcare experiences of families affected by Huntington disease: need for improved care, Chronic Illn. 2011, 20; 7: 3–225.

42. Dharssi S., Wong-Rieger D., Harold M. and Terry S., Review of 11 national policies for rare diseases in the context of key patient needs Orphanet Journal of Rare Diseases. 2017;12:63.

43. Baumbusch J., Mayer S., Sloan-Yip I, Alone in a Crowd? Parents of Children with Rare Diseases’ Experiences of Navigating the Healthcare System, Journal of Genetic Counseling. 2019; 28: 80–90.

44. Anderson M., Elliott EJ., Zurynski YA.. Australian families living with rare disease: experiences of diagnosis, health services use and needs for psychosocial support. 2013. Orphanet J Rare Dis; 8(1):22.

45. Forsythe LP., Szydlowski V., Murad MH., Ip S., Wang Z., Elraiyah TA., A systematic review of approaches for en-gaging patients for research on rare diseases, Journal of General Internal Medicine, 2014; 29(S3), 788–800.

46. Grut L. & Kvam MH., Facing ignorance: people with rare disor-ders and their experiences with public health and welfare services. Scandinavian Journal of Disability Research, 2013; 15(1), 20–32.

47. Huyard C., What, if anything, is specific about having a rare disorder? Patients’ judgements on being ill and being rare. Health Expectations, 2009; 12(4), 361–370.

48. Dellve L., Samuelsson L., Tallborn A., Fasth A., & Hallberg LRM., Stress and well-being among parents of children with rare diseases: a prospective intervention study, Journal of Advanced Nursing, 2006; 53(4) 392–402.

49. Farmer JE., Marien WE., Clark MJ., Sherman A., & Selva TJ., Primary care supports for children with chronic health conditions: identifying and predicting unmet family needs. Journal of Pediatric Psychology. 2004; 29(5), 355–367.

50. Taruscio D., Arriola L., Baldi F., Barisic I., Bermejo-Sanchez E., Bianchi F., Calzolari E., Carbone P., Curran R., Garne E., Gatt M., Latos-Bielenska A., Khoshnood B., Irgens L., Mantovani A., Martinez-Frias ML., Neville A, Ribmann A., Ruggeri S., Wellesley D., Dolk D., European Recommendations for Primary Prevention of Congenital Anomalies: A Joined Effort of EUROCAT and EUROPLAN Projects to Facilitate Inclusion of This Topic in the National Rare Disease Plans, Public Health Genomics. 2014; 17: 115–123.

51. Rodwell C. and Ayme S., Rare disease policies to improve care for patients in Europe, Biochimicia Biophys Acta. 2015;1852 (10Pt B):2329–35.

52. Wheelwright S., Darlington A-S., Hopkinson J.B., Fitzsimmons D. and Johnson C., A systematic review and thematic synthesis of quality of life in the informal carers of cancer patients with cachexia, Palliative Medicine, 2015; 30; 2.

53. Santin O, McShane T., Hudson P. and Prue G., Using a six-step co-design model to develop and test a peer-led web-based resource (PLWR) to support informal carers of cancer patients, Psycho-Oncology, 2018; 28; 3.

54. Katende G., Nakimera L., (2017), Prevalence and correlates of anxiety and depression among family carers of cancer patients in a cancer care and treatment facility in Uganda: a cross-sectional study, 17; 3.

55. Strauss AL and Corbin J. Basics of Qualitative Research, 3^rd^ edition, London Sage, 2008. p31–34

56. SmartSurvey. www.smartsurvey.co.uk. Accessed 06 May 2020.

57. Queen’s University, Belfast, Rare Disease webpage. https://www.qub.ac.uk/sites/RareDisease/ Accessed 28 April 2020.

58. Bryman A. Social Research Methods, 4^th^ Edition, Oxford, Oxford University Press, 2012. p405–406.

59. Ritchie J, Lewis J, McNaughton, Nicholls C and Ormston R. (eds) Qualitative Research Practice - A Guide for Social Science Students and Researchers, London, Sage Publications. 2014 p4–6.

60. Marshall C. and Rossman GB. Designing Qualitative Research, 6^th^ Edition, London, Sage Publications Ltd, 2016. p147–155.

61. NVIVO https://www.qsrinternational.com/nvivo-qualitative-data-analysis-software/home Accessed 1 May 2020.

62. Silverman D. Interpreting Qualitative Data, 3^rd^ Edition, London, Sage, 2006. p76–78

63. McQueen R., and Knussen C. Research Methods for Social Science, An introduction, Essex, Prentice Hall, 2002, p200–201.

64. Wertz FJ., Charmaz K., McMullen L.M., Josselson R., Anderson R. and McSpadden E. Five Ways of Doing Qualitative Analysis, New York, The Guillford Press, 2011, p58–59.

